# Application of Carleman approximants for the estimation of epidemic parameters from incidence data time-series

**DOI:** 10.1101/2022.07.14.22277622

**Authors:** Juan C. Muñoz-Sánchez, Santiago F. Elena, José–Angel Oteo

## Abstract

We have analyzed the possibility of estimating epidemiological parameters from daily infection incidence data. In particular, we have focused on the determination of the instantaneous reproduction number, the contagion period and the duration of the infectious period using only the reported incidence time-series information. We have developed a data-driven method based on the instantaneous mapping of the infection incidence data on the simplest (two parameter) SIR model, along the progression of an epidemy. The mapping is carried out via Carleman linearization of the non-linear model equations. We concluded that the daily infection incidence series on its own does not carry enough information to provide estimates for the above time scales and hence additional measurements and/or hypotheses must be considered. In contrast, the prevalence time-series does allow for accurate estimates. For the case in which the characteristic infectious period is available, a new algebraic formula for the instantaneous reproduction number has been derived.

## 1 Introduction

The evolution of an epidemic in the course of time may be, ideally, characterized by a limited number of dynamical varying parameters. Mathematical models provide a way to define them in a precise manner [1, 5, 15, 18, 21, 28]. Their values have to be estimated from available observational data, a not always simple task. Daily incidence data are time-series readily accessible and expected to contain dynamical information in amplitudes as well as in statistical correlations. Here we address the question of what relevant information does carry the daily incidence time-series on its own about parameters driving the epidemic process.

The most elementary epidemic model one can think of must deal with two different characteristic time scales: (*i*) the time period that an infected individual remains able to infect other individuals, and (*ii*) the characteristic time period between contagions. The interplay between their relative magnitudes determines the nature of the epidemic behaviour. One specific goal of the present study is to explore to what extent the epidemic daily incidence time-series carries enough information to ascertain both time scales.

The characteristic time scales of an epidemic process may change as it unfolds, *e*.*g*., due to changing social distance measures or the way isolation of infected individuals is implemented. The idea we develop here consists in mapping short time lapses of the incidence curve onto the simplest (two parameter) Susceptible-Infected-Recovered (SIR) deterministic model whose variables and parameters have a precise and well-defined meaning [18, 21, 22]. Thus, the hypothesis we handle is that the SIR model may provide a good short-time approximation to the epidemic incidence.

The determination of the infectious period and the contagion period are active research subjects on their own [2, 6, 11, 13, 20, 25, 26, 36]. Both of them are essentially treated from a probabilistic viewpoint. Estimating the infectious period is of great public health relevance but is difficult and the number of statistical methods described is not large [36]. The contagion period is closely related to the so-called generation time which is the time from being infected to generate a secondary infection; although in practice, the so-called serial interval is used instead [12]. It refers to the time from illness onset in the primary case to illness onset in the secondary case, and illness onset refers to symptom onset time. The common formulae [4, 10, 14, 24, 29, 30, 32, 35] to compute the instantaneous reproduction number *R*_*t*_, an important epidemiological parameter to be defined below, are based on serial interval estimates. To this respect, we derive a new algebraic formula to estimate *R*_*t*_ on the basis of the infectious period duration. As thoroughly discussed by Vegvari et al. [33], the use of deterministic dynamical models is one approach among others. Incorporating stochastic assumptions in the model, agent-dependent models, statistical models, or phylogenetic methods are alternative frameworks. We will compare our results with those from statistical models, which are the most common empirical methods to estimate *R*_*t*_.

The main conclusion we have reached is that the daily epidemic incidence data series, expressed as a population fraction (say *i*(*t*)), does not carry on its own enough information to allow precise estimates of these time scales. At least when it is piecewise mapped onto the SIR model. In contrast, in combination with the prevalence (or population infective fraction time-series), say *I*(*t*), which is a sort of aggregate daily incidence series, such estimates are allowed. Indeed, *I*(*t*) may be thought as the sum (possibly weighted) of previous daily incidence data along a number of *a priori* unknown days. Note that this drawback does not stem from the many artifacts that plague the daily reports of observed incidence data series. The key problem is that *I*(*t*) is a native variable of the SIR model whereas the measured quantity *i*(*t*) is not. The relationship between both variables conveys, at least, knowledge of the characteristic infectious period. This point turns out to be crucial in a data-driven determination of epidemic parameters.

### 1.1 Fitting the dynamical model to observed data

The procedure is as follows. We consider the two time-series quoted above: the daily incidence fraction, *i*(*t*), and the infective fraction *I*(*t*) or prevalence. The latter, together with the fractions of susceptible (*S*(*t*)) and recovered (*R*(*t*)) population categories at time *t* constitute the dependent variables of the epidemiological SIR model

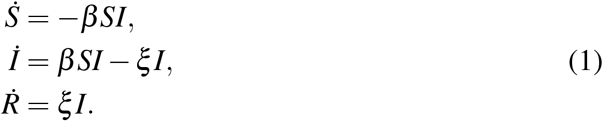

Here the dot stands for time derivative and because *S*(*t*)+*I*(*t*)+*R*(*t*) = 1, the population is constant. This is an initial value problem to be solved numerically from initial conditions: *S*(*t*_0_) = *a, I*(*t*_0_) = *b,R*(*t*_0_) = 1 − *a* − *b*. The SIR model is among the simplest one of the wide family of compartmental models used in infectious disease epidemiology. It is the simplest option in the sense that three degrees of freedom is the smallest phase space dimension allowing a solution *I*(*t*) exhibiting a maximum, namely a true epidemic wave. The population size is assumed to stay constant, which amounts to say that the effect of births and deaths is discarded. This approximation is consistent with an application of the model to short enough time intervals.

The approach we follow is to model an epidemic curve by piece-wise SIR models. Every particular SIR model is then characterized by slightly different values of the parameters *β* and/or *ξ*. The concatenation of all of them in short time lapses is intended to reproduce the epidemic curve. Our goal is to solve the inverse problem: given the outbreak incidence curve we aim to determine the succession of SIR parameter values that better accounts for it. In this context, the use of more sophisticated compartmental models would be justified only if long enough fitting time intervals were used, so that the new variables become relevant.

The dimensions of the parameters *β* and *ξ* are inverse of time. Thus, they are rates associated with the characteristic time scales of the system. In particular, 1*/β* stands for the characteristic time scale at which contagion takes place and 1*/ξ* is the characteristic time during which an infective subject can infect. Equivalently, *β* corresponds to the contagion rate and *ξ* to the recovery rate. Social distance measures act on the *β* value. The way infected individuals are detected and isolation measures implemented affect the *ξ* value. We could even argue, *e*.*g*., that different lineages of the same pathogen could have different infectious periods.

### 1.2 The instantaneous reproduction number and beyond

The definition of the basic reproduction ratio, *R*_0_, in the SIR model reads *R*_0_ ≡ *β/ξ*. It is the particular case of the instantaneous reproduction ratio quoted above, *R*_*t*_ = *R*_0_*S*(*t*), at the beginning of the outbreak because *S*(0) = 1. This definition may be reworded as the average number of new infections caused by a single infected individual at time *t* in a partially susceptible population. The information provided by *R*_*t*_, is important. For instance, according to (1), *İ*= *ξ* (*R*_*t*_ − 1)*I*, and whenever *R*_*t*_ > 1 the prevalence *I*(*t*) increases. However, the answer to other questions requires the knowledge of either both time scales or further parameters. To motivate this issue, consider the relationship between *I*(*t*) and *i*(*t*). The epidemic incidence in the SIR model is given by *i*(*t*) = *β S*(*t*)*I*(*t*)Δ*t*, where Δ*t* stands for the sampling time interval under consideration. Thus, if Δ*t* = 1 day, then *β* and *ξ* are measured in days^*−*1^. A possible connection between the two time-series *I* and *i*, requires the explicit knowledge of *β* and *S*(*t*). In a second instance, the value of the infective fraction *I*(*t*) should be an aggregate of *i*(*t*) values along a number of days that should be ruled by the infectious period 1*/ξ*, as we have already pointed out.

Another example is to forecast the inflection point of a growing outbreak, namely to ask for a change in trend from acceleration to deceleration. An answer to this question involves, besides *R*_*t*_, the fraction *I/S*. To see this, we look at the second derivative sign of *I* which determines the type of growth

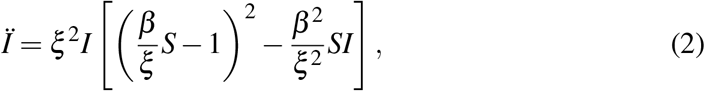

and leads eventually to the discriminant condition

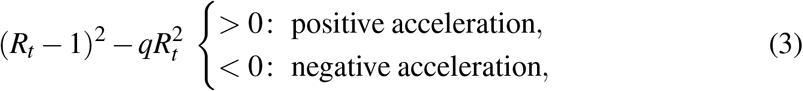

where, with similar notation as above, *S*(*t*) = *a, I*(*t*) = *b* and *q* ≡ *b/a* at the time *t* under scrutiny. For the worrisome case *R*_*t*_ > 1, the condition (3) reduces to

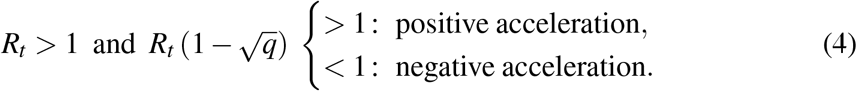

This result shows how the time of the inflection point depends concomitantly on both *R*_*t*_ and *q*.

All these examples point out that the knowledge of the parameter *R*_*t*_ is not enough to fully characterize the dynamical behaviour of an epidemic and eventually to forecast its evolution. Thus, estimates of the two time scales 1*/β*, 1*/ξ*, and the relative fraction of infected/susceptible *q* are relevant too.

### 1.3 The goal

The procedure we develop in the next sections is intended to provide estimates for all four parameters *ξ, β, a, b*, at time *t*, from two different viewpoints. Firstly, we consider as input data the values *I*(*t*) obtained from numerical simulations. The incidence is then *i*(*t*) = *β S*(*t*)*I*(*t*)Δ*t*. In this situation the method provides good parameter estimates. Secondly, we carry out a similar analysis with *i*(*t*) as input data. In this case, the values *S* and *I* from the numerical integration are used just to build *i*(*t*). This corresponds to the situation encountered when analyzing real data. The prevalence, when needed, is estimated from *i*(*t*). It is shown that the information in *i*(*t*) is incomplete to this end.

The fit of the SIR system (1) to epidemiological observed data may be a non-trivial numerical task. To facilitate it, we have introduced the Carleman linearization method [3] for the first time to this problem. The idea is to replace the non-linear differential equations system (1) with an approximate linear one of higher dimension which is easily solvable analytically. We use then the algebraic approximate solution to carry out the non-linear fit to epidemiological data series. The use of Carleman instead of polynomial approximants improves the quality of the approximate algebraic SIR solutions involved in the fits and, besides, allows us to build up an algebraic approximate formula to estimate *R*_*t*_.

In Section 2, the Carleman linearization scheme is explained. In Section 3, the two types of simulations are carried out where the input data are alternatively prevalence or incidence series. Results from real epidemiological data are given in Section 4. In Section 5, an algebraic formula for *R*_*t*_ is derived and discussed.

## 2 Carleman linearization of the SIR model

The idea behind the Carleman linearization procedure [3, 7, 23, 27] consists of repeatedly replacing non-linear terms in (1) with new linear dependent variables at the price of adding new non-linear differential equations to the system. Ideally, the non-linear system (1) is then equivalent to an infinite dimensional linear one. We will handle a linear truncated version that is expected to provide a good analytical approximation of the solution in an appropriate time interval. Next, we illustrate the first two replicas of the procedure.

We begin by replacing *f* = *SI* in (1) and providing the differential equation for *f*.

Without loss of generality we shift the origin (*t* = 0) to the point of interest

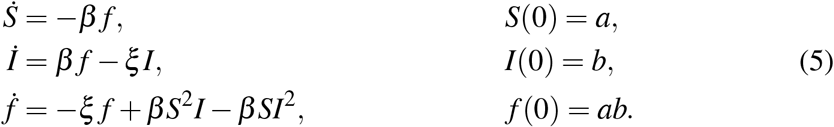

Notice that the differential equation for the recovered fraction *R*(*t*) in (1) is decoupled and therefore has been left aside in the procedure. The system of two first equations in (1) and the system (5) are mathematically equivalent. Removing the two *β* Terms in the right-hand side of the third equation yields a linear differential system whose solution provides the first closed form approximate solution to the original system

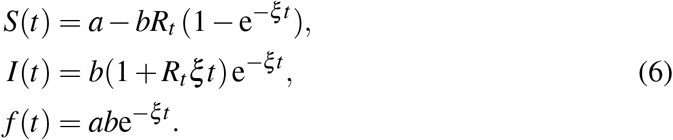

The procedure can be replicated with the replacements *g* = *IS*^2^ and *h* = *SI*^2^ to get

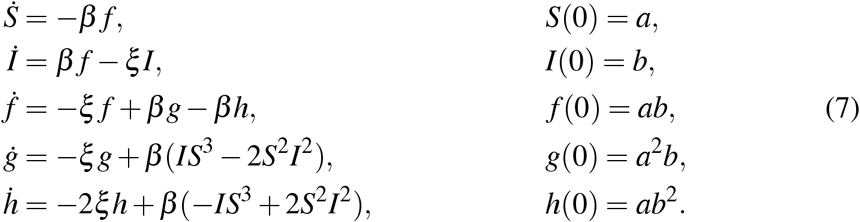

If we drop out the *β* terms in the last two equations, the system becomes linear and provides the second order analytical approximate solution. For the SIR system the procedure can be easily transformed into a recurrence for higher orders of approximation. The coefficient matrix of the linear differential system is triangular and therefore the solution is easily obtained.

The leading term in the solution of *I*(*t*) is exponential-like, a fact witnessed in the approximation (6). For this reason, we have introduced a change of variable, namely *I* = e^*Y*^, in the SIR system prior to the Carleman linearization of (1). As a consequence, the equation *İ* = *β f − ξ I* is replaced with 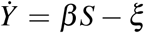, *Y* (0) = ln *b*. The resulting system of Carleman differential equations is then non-homogeneous but its resolution is not much more involved. Had we kept the full system of infinite dimension, this change of variable would be irrelevant. However, as a consequence of the truncation, every exponential approximation e^*Y*^ keeps an infinity of powers *β*^*n*^, unlike the original variant *I*. This change of variable can be interpreted as a kind of re-summation technique. Hereafter, we will refer to this variant as the SYR system.

To follow the exposition bellow, and to derive the algebraic formula for *R*_*t*_, it is useful to keep in mind the particular solution (6) or its corresponding SYR version

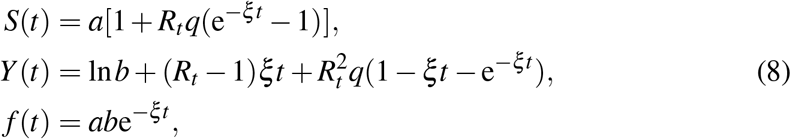

albeit the numerical fits have been done with higher order approximations (see Appendix for the second and third order approximants).

## 3 Simulated data

We have carried out a number of analyses corresponding to several epidemiological settings using numerical simulations. First of all, in order to assess the utility of Carleman analytical approximants we illustrate in Figure 1 the behaviour of some of them for different approximation orders. The curves stand for an epidemic wave generated using the SIR model with fixed parameter values. For fixed order of approximation, the local behaviour with these approximations is better than with the corresponding polynomial approximants (not shown in the Figure 1). Expressions for polynomial approximants may be obtained by truncating the Taylor expansion of Carleman ones. The idea of working with the more involved Carleman algebraic ansatz is to improve as much as possible the fitting part of the procedure. Here the fifth order of Carleman approximation has been used.

**Figure 1:**
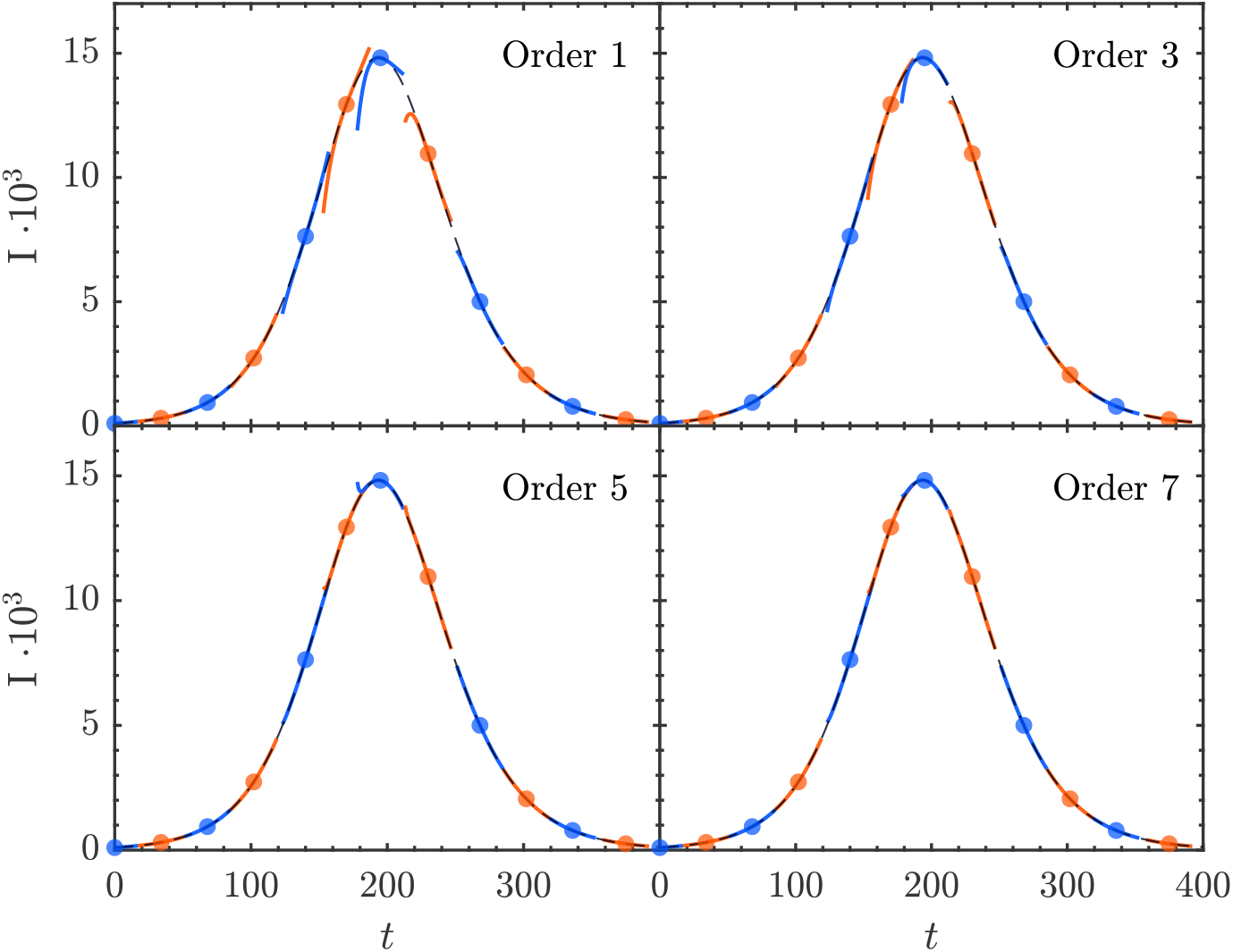
Carleman analytic approximants for the prevalence *I*(*t*) of a one-wave epidemic generated using the SIR model with fixed parameter values *ξ* = 1*/*6, *β* = 1*/*5. The approximation order is indicated in every panel.

Two classes of simulation have been performed. In the first case, we use as input the SIR prevalence fraction values {*I*_*k*_} obtained by numerical integration of (1) and the daily incidence fraction values, computed as *i*_*k*_ = *β I*_*k*_*S*_*k*_Δ*t*. From the epidemiological data availability viewpoint, the knowledge of {*I*_*k*_} is not the common situation. It corresponds, if any, to outbreaks in very small populations where onset and end of symptoms (taken as a proxy of infectiveness) for every subject are recorded.

In the second case, we take as input {*i*_*k*_} computed from the numerical solutions *I, S*. After that, the infective fraction {*I*_*k*_} used in this simulation is reconstructed using two different schemes. Later, some noise is added to the synthetic data series to gain realism. This simulation is intended to mimic the common situation where the observed data {*i*_*k*_} are the only input.

The reconstruction of *I* from *i* is certainly a drawback because the explicit connection between the true SIR variable *I* and the non-SIR variable *i* is a problem in itself. We have figured out two very different ways intended to render the conclusions independent, as far as possible, of the particular details used in the *I* reconstruction. In the first one, it is assumed that an infective subject infects only during a *D* days period, therefore

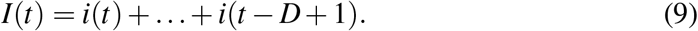

Alternatively, a discretized version of (1) reads: *I*(*t*) − *I*(*t* − 1) = [*βI*(*t* − 1)*S*(*t* − 1) *ξI*(*t* − 1)Δ*t* = *i*(*t* − 1) *ξI*(*t* − 1) Δ*t* and yields the recurrence *I*(*t*) = *i*(*t* − 1)+(1 − Δ*t/D*)*I*(*t* 1), with 1*/D* the intended true value for *ξ*. The recurrence solution reads

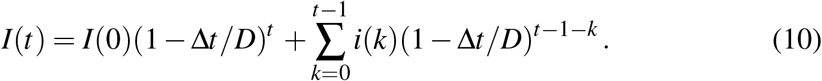

Both alternatives may be thought as weighted sums of past incidence values.

The question about what is the most convenient cost function as regards the non-linear Least Squares (LS) fits of the time-series is shared by both simulation classes. The straightforward approach, because of the direct availability of data, is to consider the LS fit to daily incidence data, say {*i*_*k*_}. The daily incidence in the model is given by *β f* (*t*), and the cost function *χ*^2^ reads then

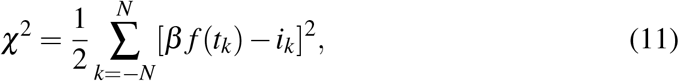

with *f* (*t*) given by a Carleman approximant. Besides the non-linearity of the variational equations, the drawback in this scheme is that the leading term in *β f* (*t*) is ruled by the threefold parameter product *βab* (see, *e*.*g*., (6)), which is a hindrance for the minimization algorithm. To avoid it, we choose to fit both incidence and (log of) prevalence with the cost function

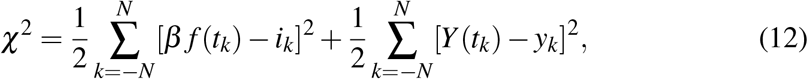

where *y*_*k*_ ≡ ln(*I*_*k*_). Here 2*N* + 1 stands for the number of points in the time interval of the fit.

The non-linear LS minimization has been performed using the Matlab routine *lsqcurvefit* [19].

### 3.1 First simulation class: *I*(*t*) as input

In Figure 2 we have represented a two-wave outbreak obtained by integrating the model equations (1) with constant *ξ* = 1*/*5 and varying *β*. We have added some amount of noise to the solution *I*(*t*). The daily incidence has been reconstructed as *β IS*Δ*t*, with Δ*t* = 1 day, from the numerically integrated solutions.

**Figure 2:**
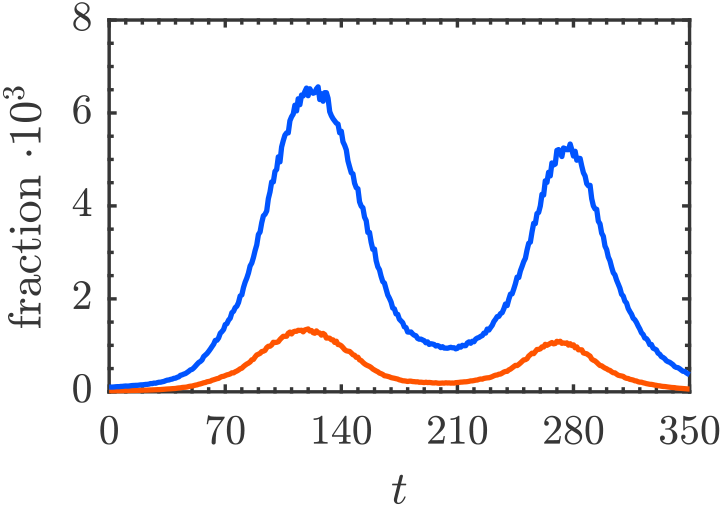
Two-wave SIR epidemic with added noise used in the simulation. *ξ* = 1*/*5 and varying *β*. Infective fraction *I*(*t*) (blue) and daily incidence fraction *i*(*t*) (red) as a function of time.

The estimated parameters are plotted in Figure 3 as well as their corresponding values used for the numerical simulation. The *R*_*t*_ nominal value of the model (dashed line) corresponding to the numerical solution and the outcome of the fit is set out in Figure 3A. With same plot coding, we give in Figure 3B the *q* ratio, namely, the fraction infective over susceptible. Eventually, we plot in Figure 3C the estimates for *ξ* and in Figure 3D for *β*. Clearly, the estimates are reasonable. This pattern replicates in other simulations we have carried out, for instance with varying *ξ* and fixed *β*. This is to say that whenever the analysis proceed from the prevalence *I* as input data, the parameter determination achieved is correct.

**Figure 3:**
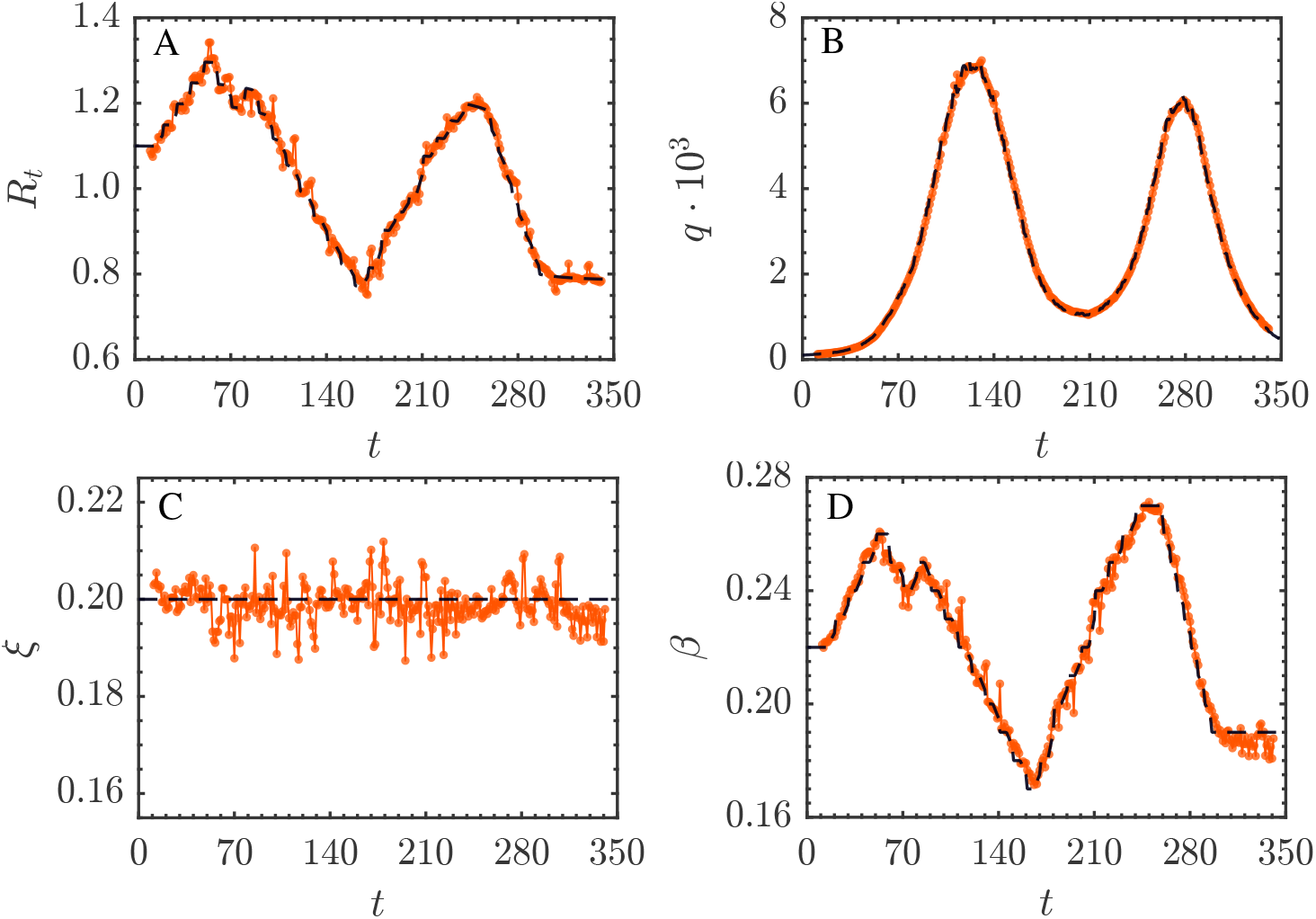
Two-wave SIR first class simulation outcomes. Dashed lines stand for nominal values.

### 3.2 Second simulation class: *i*(*t*) as input

We have replicated the analysis above using the daily incidence *i*(*t*) as input data. The prevalence *I* has then been built up using the two methods (9) and (10). The panels in

Figure 4 show the respective outcomes for *R*_*t*_, *q, β, ξ*. The different lines correspond to a number of choices for *D* and are to be compared with the nominal result.

**Figure 4:**
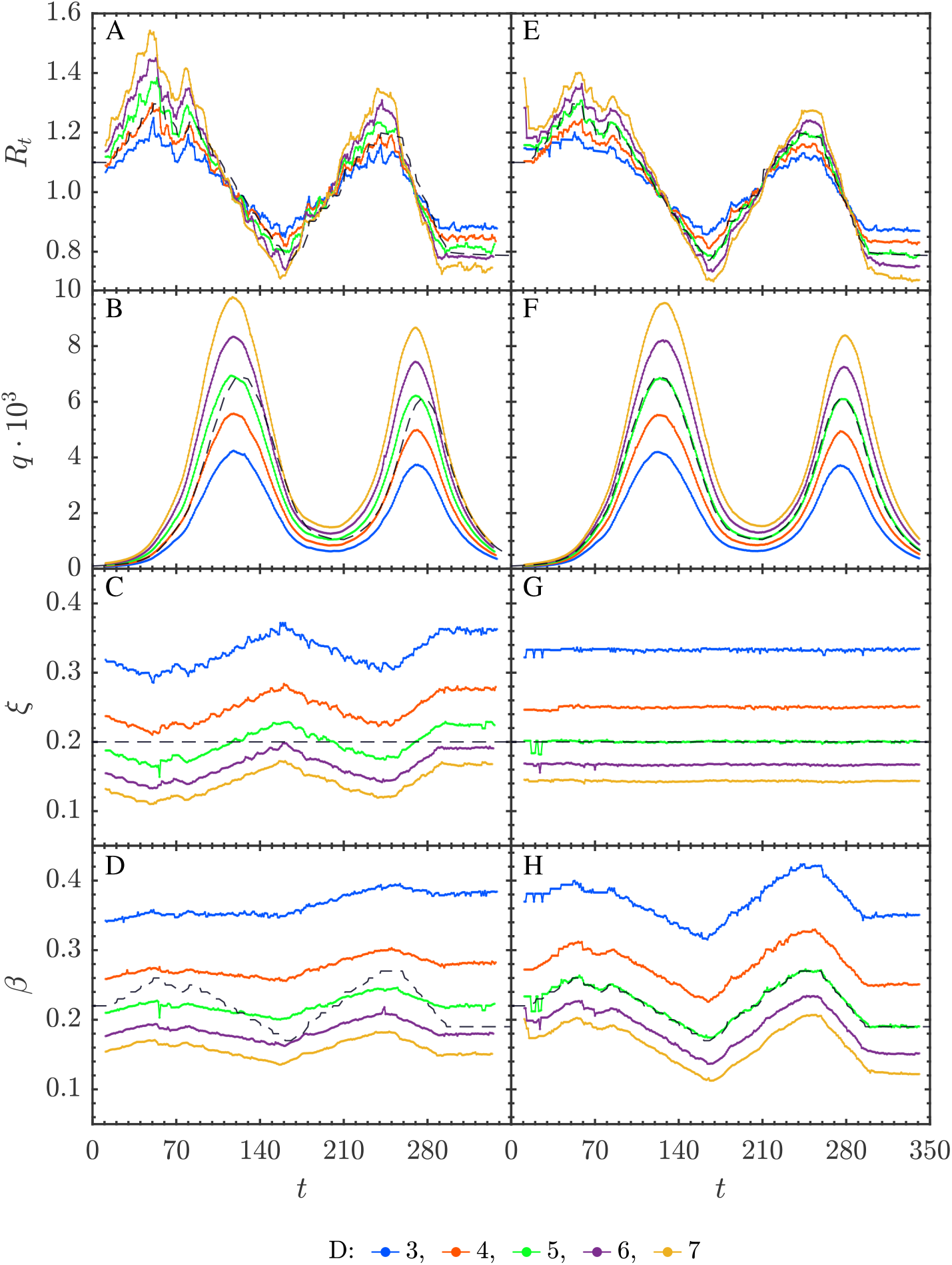
Two-wave SIR second class simulation with prevalence reconstruction using (9) for A to D panels and (10) for E to H panels. Dashed lines stand for nominal values.

It is clearly seen that all the outcomes are sensitive to the *D* value used in (9) and (10). The most conclusive result is in Figure 4G where the *ξ* value recovered after the fit is just the very value 1*/D* used to reconstruct *I*(*t*) from *i*(*t*). The corresponding panel in Figure 4C exhibits oscillating patterns around the value 1*/D*. The variability in the outputs for *q, β, ξ* is large, and milder in the *R*_*t*_ outcome. Notice that this simulation class mimics a situation with field data.

## 4 Fits to epidemiological incidence data

To explore the behaviour of the method on field data we choose to fit epidemiological incidence data of SARS-CoV-2 provided by the Government of Valencian Community (CV), Spain [17]. Figure 5 outcomes the reported daily cases, as well as the official *R*_*t*_ estimate as a function of time [16]. This situation compares to the numerical simulations of the second class above.

**Figure 5:**
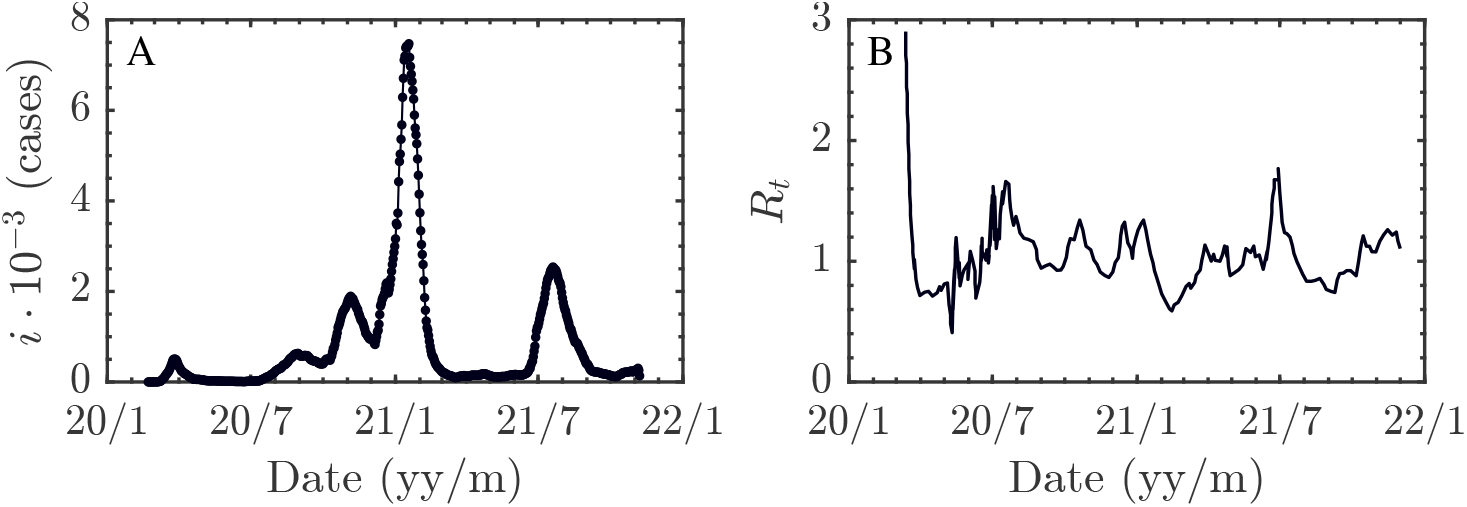
A) SARS-CoV-2 reported daily cases for the CV [17] and B) the corresponding official *R*_*t*_ estimate [16].

Figure 6 illustrate *R*_*t*_, *q, ξ* and *β*. Every line corresponds to a different *D* value in the prevalence reconstruction from the incidence data. For both reconstruction methods, the *R*_*t*_ estimates remain quite stable with respect to the *D* value along the time. However, the *ξ* estimates exhibit the very same behavior described in simulations (see Figures 4C and 4G). Namely, the *ξ* outcomes of the fit tend to approximate 1*/D*: the input used in the prevalence data reconstruction.

**Figure 6:**
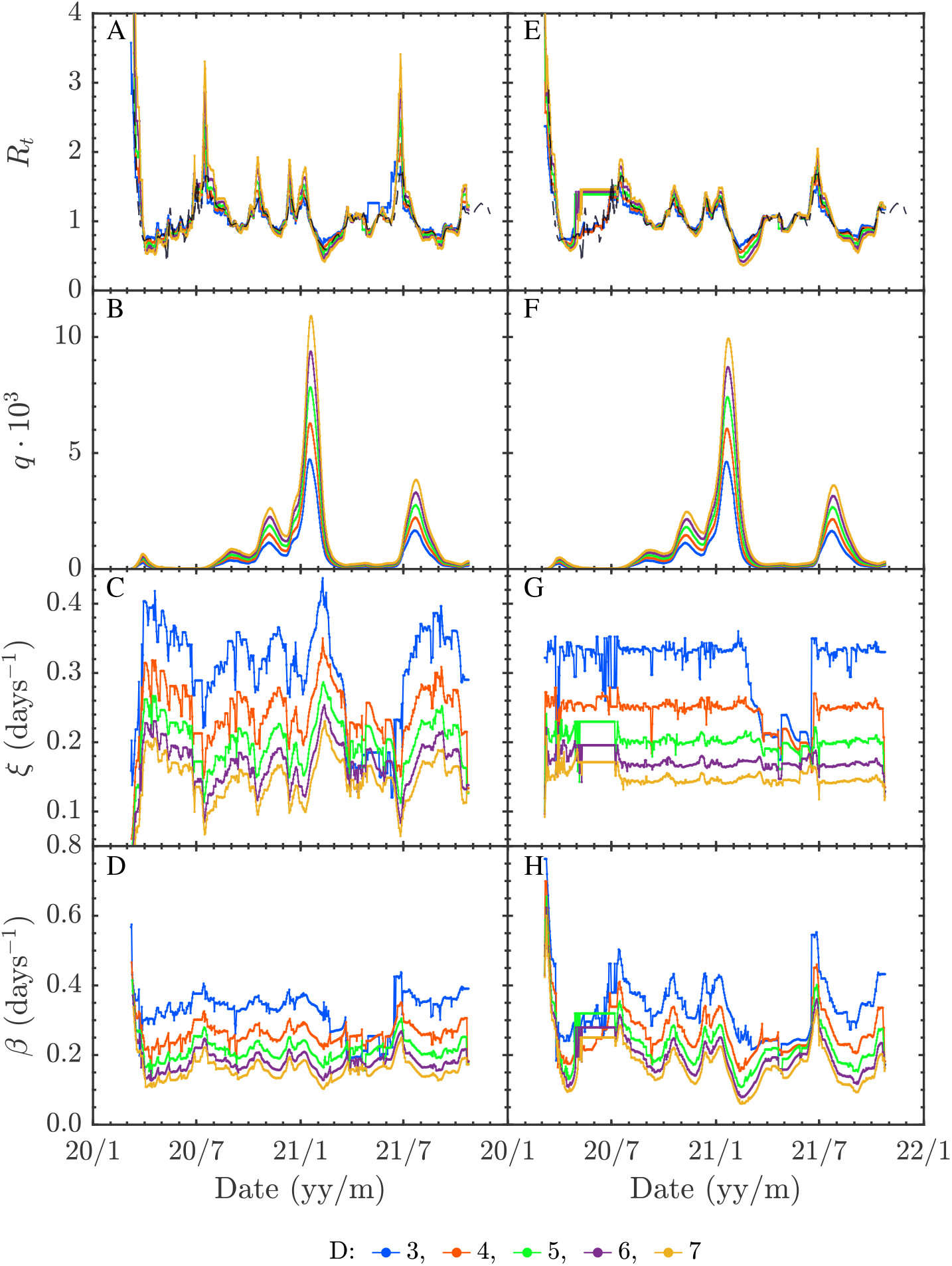
CV data estimates. *I*(*t*) obtained from *i*(*t*) according to (9) for A to D panels and (10) for E to H panels. Dashed line in panels A and E stand for the official *R*_*t*_ estimate.

## 5 An *R*_*t*_ formula when the infectious period is known

If we assume that (*i*) the infectious period 1*/ξ* is known by any alternative method and, (*ii*) a reliable method to reconstruct the prevalence *I* from the incidence *i* has been implemented, then some simple algebraic formulas can be obtained from the framework we have developed. We proceed with the Carleman approximant (8) and minimize

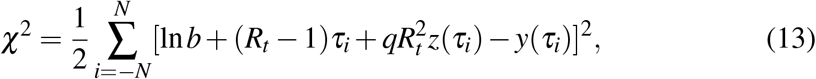

with respect to ln *b, R*_*t*_, and *q*. We have defined *τ* ≡ *ξt*, and *z*(*τ*) ≡ 1 − *τ* − exp (−*τ*). Remind that *y*(*τ*_*k*_) = ln(*I*(*τ*_*k*_)). The resulting variational equations are solvable in closed form and the algebraic solution reads

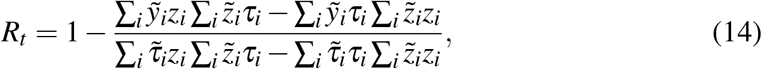

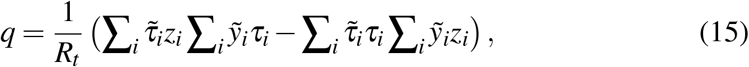

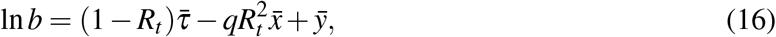

in terms of the following quantities

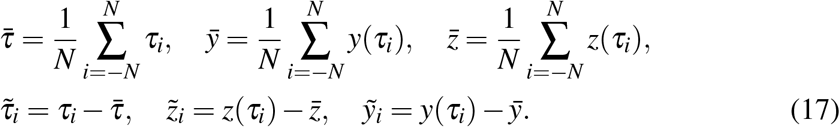

The solution (16) comes directly from 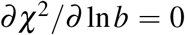 whereas 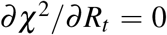and 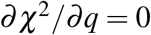can be simplified to the coupled system

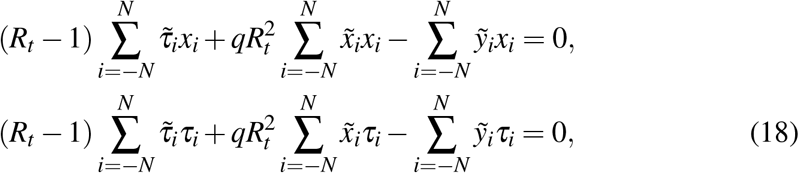

which yields (14) and (15). Notice that the solutions come in terms of the *dimensionless time τ*, not *t*, which tells us about the crucial role of *ξ* describing the correct time scale of the process.

Even though the Carleman approximant (8) is an elementary approximation to the analytical SYR solution, it turns out that equation (14) yields estimates in good agreement with both numerical simulations and CV data reports as we show next. However, at odds with this, the performance of equations (16) and (15) is not accurate enough. For this reason, we focus our discussion just on the *R*_*t*_ formula. To this end, we define the score

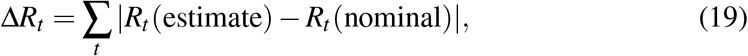

where the sum goes along the full time interval analysed. Figures 8 and 9 outcome (14) and (19) for several situations:

1. Figure 8. Simulations from prevalence *I*(*t*) data with several *D* values used in (9) and (10) and fixed fit range: *N* = 6 days. To choose the *N* value we have carried out a number of simulations (see Figure 7) with 1 ≤ *N* ≤ 12. Figure 7B shows the score obtained vs. *N* and points out that *N* = 6 is optimal. The behaviour of *R*_*t*_ as a function of *D* is biased as in the simulations of Figure 4. The score (19) in Figure 8B confirms the value *D* = 5, which is the one used in generating the numerical *I*(*t*) values.
2. Figure 9. Simulations from incidence *i*(*t*) data with several *D* values used in (9) and (10) and fixed fit interval: *N* = 6 days, using the two prevalence reconstruction schemes (9) and (10), Figures 9A and 9B. This simulations mimic a field data analysis case. The *R*_*t*_ estimates are biased as in all the preceding cases. This conclusion holds for both prevalence reconstruction methods. Therefore, the crucial point is not the way followed to obtain *I*(*t*) from *i*(*t*) but to handle the correct estimate for the infectious period duration.

**Figure 7:**
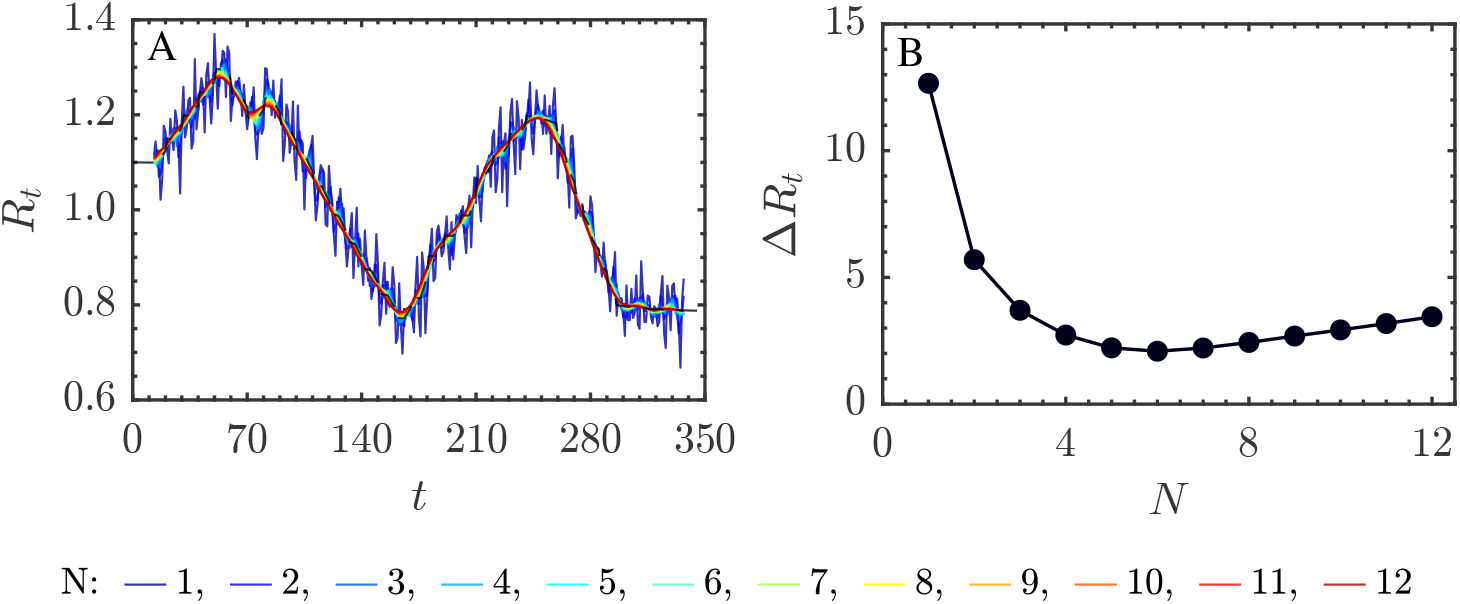
A) *R*_*t*_ estimate using *I*(*t*) data and the analytical approximation (14) for a number of fit ranges (*N*) and fixed infectious period 1*/ξ* = 5 days. B) Δ*R*_*t*_ score (19). According to the flat shape of the minimum the conclusion is that a value around *N* = 6 gives the appropriate fitting range.

**Figure 8:**
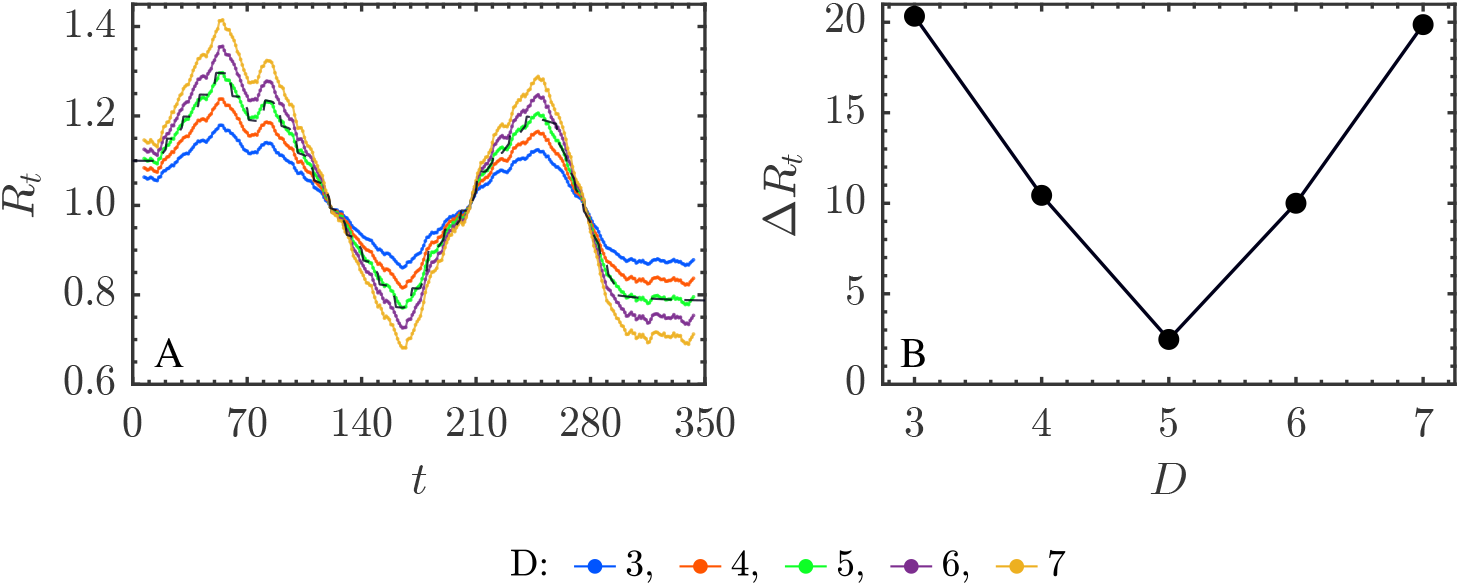
A) *R*_*t*_ estimates using simulated data and the analytical approximation (14) for different values of *D* and fixed interval *N* = 6 in (14) and B) the score Δ*R*_*t*_ in (19).

**Figure 9:**
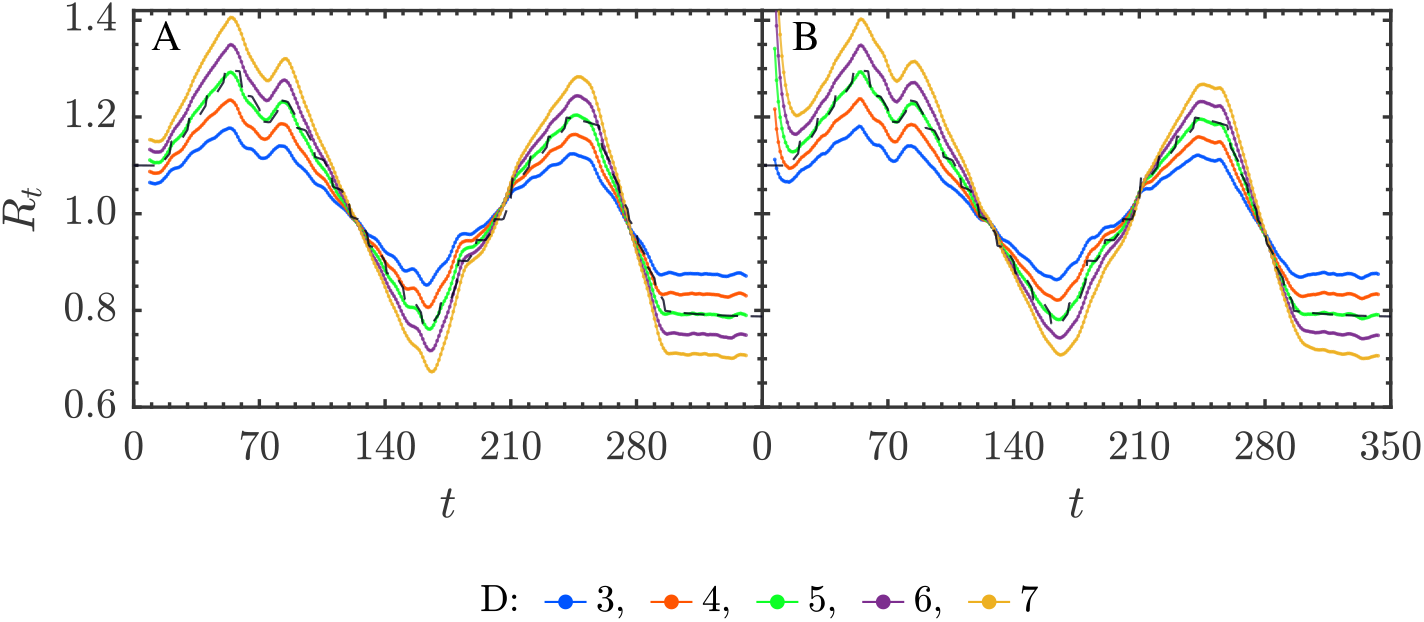
*R*_*t*_ estimates obtained according to (9), in (A) and (10), in (B), using (14) for different *D* values and *N* = 6. In both situations the score Δ*R*_*t*_ is similar to the one in Figure 8B with a minimum at *D* = 5 days.

Eventually, we have checked (14) with the SARS-CoV-2 CV daily incidence data in Figure 5. Figure 10A and 10B illustrate the *D* dependent bias of *R*_*t*_, which is similar to that observed with the numerical simulations.

**Figure 10:**
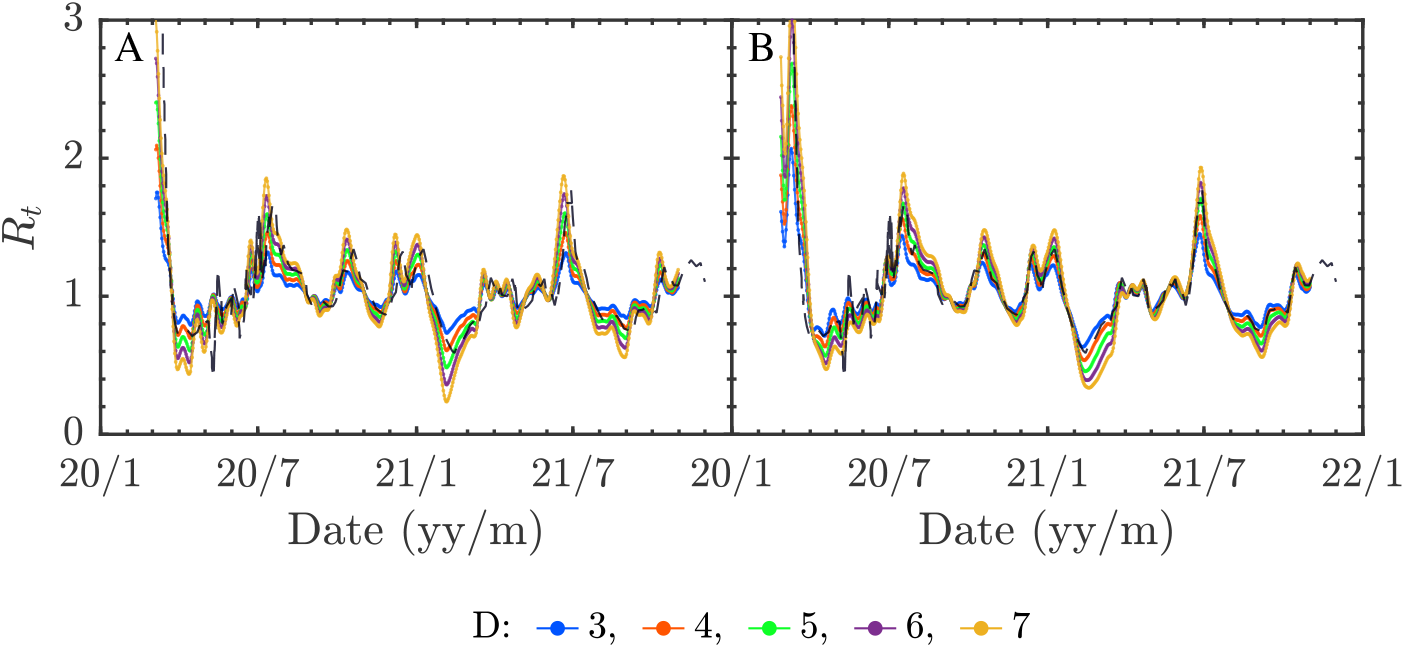
*R*_*t*_ estimates using (14) for CV data. The reconstruction of *I*(*t*) has been carried out with (9) in (A) and (10) in (B). Fit range *N* = 6 and different *D* values. Dashed line stands for the official *R*_*t*_ estimate.

## 6 Discussion

In this study we have raised the question of whether the information contained in the epidemic incidence time-series by itself (in form of amplitudes and time correlations) would be enough to allow the determination of reliable estimates of epidemiological parameters. Our answer is no, at least as it regards to estimating the instantaneous reproduction number, the infectious period and the contagion period.

We have piece-wise mapped the epidemic incidence onto the simple SIR model where the quantities referred to above are well defined. The key point is that the native variable in the SIR model is the prevalence and not the incidence. As a consequence, the estimates for the prevalence have to be built up from the experimentally measured incidence and this procedure requires knowledge about the infectious period duration.

In the approach we have followed, the infectious period emerges as a fundamental quantity that settles the time scale of the epidemic process. The accuracy achieved for this time scale determines the estimates quality of the remaining parameters. In contrast to this, the details of the way the prevalence is estimated from the incidence seem less critical.

The framework we have developed has lead us to an algebraic formula for *R*_*t*_, provided the infectious period is known. This result, (14), is rather different from the common empirical formulae [10, 31–35] used to estimate *R*_*t*_, which are essentially based on Bayesian methods. For instance, from [10],

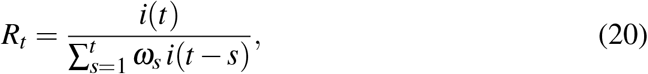

where *ω*_*s*_ stands for the probability that the interval of infection time between the primary and the secondary cases is *s*. To use this formula, the common assumption is that *ω*_*s*_ follows a discretized Gamma distribution. It is also assumed that all the infections are observed. In practice, the serial interval is used instead of the generation interval. Their distributions have the same mean but different variances, in general, which can bias *R*_*t*_ estimates [12].

The two methods (14) and (20) are rather different approaches to the same quantity, with their own pros and cons. Thus, the estimator (20) measures the infected population fraction incident on day *t* with respect to the current infective population fraction (weighted with *ω*_*s*_ the fraction that was infected *s* days in the past). The crucial information comes here from the serial, or generation, interval. These quantities are, indeed, similar to the contagion period 1*/β*. In (20), the correctness of the mean, the variance and the distribution shape of the generation time are the key point [12]. In turn, the estimated *R*_*t*_ in (14) is based on the knowledge of the infectious period and obtained from the prevalence in the interval [*t* − *N*Δ*t, t* + *N*Δ*t*], with *N* ≃ (*ξ* Δ*t*)^*−*1^. Here, it is crucial the accuracy of the infectious period estimate and, to less extent, the way the prevalence is reconstructed from the incidence.

Previous works where SIR-like models are fitted to single epidemic outbreaks can be found in the literature. *E*.*g*., in [8, 9] synthetic data and real influenza single-wave outbreaks are analysed. The framework does not correspond to piece-wise model fits, but to single-wave outbreak SIR models. The infectious period is fixed beforehand and thus the only free parameter is the contagion rate. Therefore, this situation resembles the first class simulations above.

In this study, we have assumed that all individuals became symptomatic and that the transmission is well known. Unfortunately, this is not the case for many infectious diseases, and specially for COVID-19 infected individuals, for which asymptomatic and presymptomatic transmission represent a large fraction of the total. Such biases in the daily incidence data, and neglecting the time scale of asymptomatic transmission, necessarily biases the estimates of *R*_*t*_.

Throughout this study, we have tried to answer whether daily incidence time-series is sufficient to obtain estimates of the epidemiological parameters of most interest in the course of an epidemic using the simple SIR model. In the light of our results the answer is clear: daily incidence is not enough for this task. As the native variable of this model is prevalence, we are forced to reconstruct it from incidence. This process biases the result because information on the characteristic times is required. This suggests that extra efforts are needed to determine these characteristic times dynamically throughout the pandemic. For those cases in which this information is available, we have developed a simple algebraic expression with which to obtain the instantaneous reproductive number.

## Data Availability

All data produced in the present work are contained in the manuscript

## Acknowledgements

JCMS was supported by Generalitat Valenciana Plan GenT grant ACIF/2021/296. SFE was supported by European Commission - NextGenerationEU (Regulation EU 2020/2094) through CSIC Global Health Platform (PTI Salud Global) grants SGL2021-03-009 and SGL2021-03-052. JAO work was partially supported by the Spanish Agencia Estatal de Investigación (grant number PID2019-109592GB-100).

## A Higher order Carleman approximants

### A.1 Second order approximant

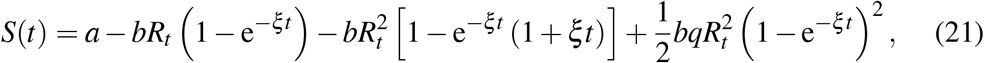

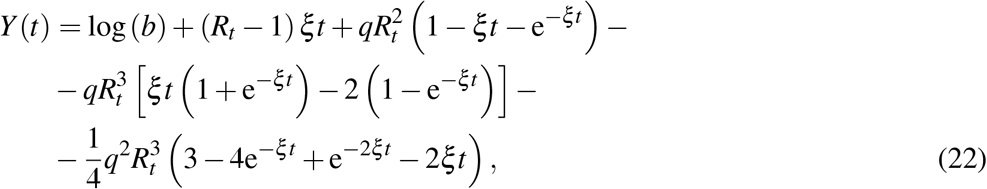

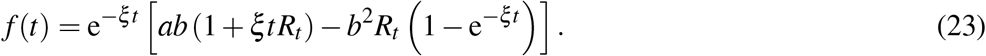

### A.2 Third order approximant

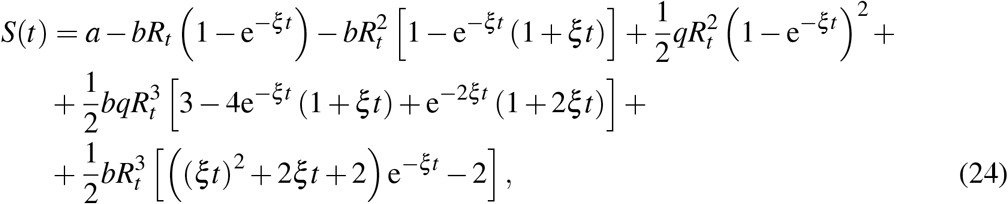

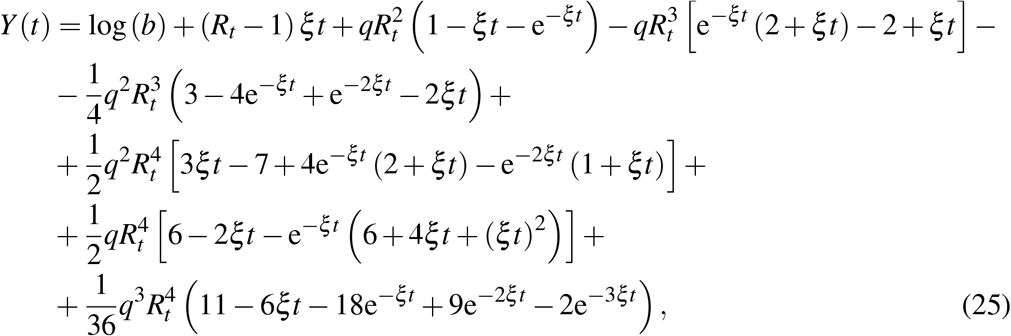

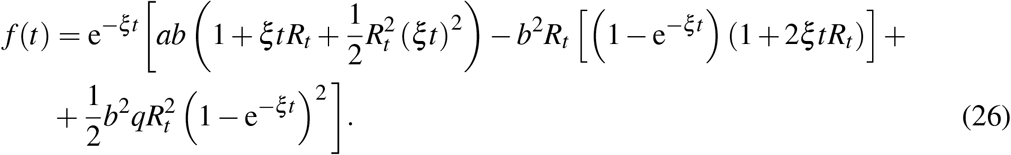

